# Determinants of the Omega-3 Index in the UK Biobank

**DOI:** 10.1101/2022.08.16.22278612

**Authors:** Jan Philipp Schuchardt, Nathan Tintle, Jason Westra, William S. Harris

## Abstract

Information on the Omega-3 Index (O3I) in the United Kingdom (UK) are scarce. The UK-Biobank (UKBB) contains data on total plasma omega-3 polyunsaturated fatty acids (n3-PUFA%) and DHA% measured by NMR. The aim of our study was to create an equation to estimate the O3I (eO3I) from these data. We first performed an interlaboratory experiment with 250 random blood samples in which the O3I was measured in erythrocytes by gas chromatography, and total n3% and DHA% were measured in plasma by NMR. The best predictor of eO3I included both DHA% and a derived metric, the total n3%-DHA%. Together these explained 65% of the variability (r=0.832, p<0.0001). We then estimated the O3I in 117,108 UKBB subjects and correlated it with demographic and lifestyle variables in multivariable adjusted models. The mean (SD) eO3I was 5.58% (2.35%) this UKBB cohort. Several predictors were significantly correlated with eO3I (all p<0.0001). In general order of impact and with directionality (- = inverse, + = direct): oily-fish consumption (+), fish oil supplement use (+), female sex (+), older age (+), alcohol use (+), smoking (-), higher waist circumference and BMI (-), lower socioeconomic status and less education (-). Only 20.5% of eO3I variability could be explained by predictors investigated, and oily-fish consumption accounted for 7.0% of that. With the availability of the eO3I in the UKBB cohort we will be in a position to link risk for a variety of diseases with this commonly-used and well-documented marker of n3-PUFA biostatus.

## Introduction

A large number of epidemiological, clinical and experimental studies have been conducted over the past few decades investigating the role of long chain omega-3 polyunsaturated fatty acids (n3 PUFAs) for health. In particular, eicosapentaenoic acid (20:5, EPA) and docosahexaenoic acid (22:6, DHA) are viewed to have beneficial effects for cardiovascular disease (CVD) ^(1–3)^, cancer ^(3,4)^, diabetes ^(5,6)^, metabolic syndrome ^(7)^, dementia or Alzheimer’s disease ^(8)^, and depression ^(9)^. However, results from randomized trials exploring the potential protective nature of n3 PUFAs on disease outcomes across this spectrum vary from beneficial to null. Because of the inability of relatively short-term (years, not decades) intervention studies to reveal the adverse effects of subclinical nutritional deficiencies, a clearer picture of the relationships between n3 PUFAs and risk for human disease may be achieved using prospective observational data based on n3 PUFA biomarkers rather than dietary intake data as the exposure.

The n3 PUFA biostatus can be assessed in a variety of lipid pools. First, n3 PUFAs can be measured across numerous blood compartments including red blood cells (RBCs), whole plasma, whole blood, platelets, leukocytes and plasma lipid classes (i.e., phospholipids, cholesteryl esters, triglycerides, and free FAs). RBCs are perhaps best suited to quantify long-term n3 PUFA blood levels since n3 PUFAs in RBCs are constant over weeks and months compared to total plasma fatty acids or plasma phospholipid fatty acids where stability is only observed over days ^(10)^. In addition, the n3 PUFA content of RBCs is similar to that of many organs including the heart, intestines, and muscle, while FAs in different compartments are less correlated with levels in other organs ^(11)^.

The Omega-3 Index (O3I) – defined as the EPA + DHA content of RBCc as a percent of total identified FAs ^(12)^ – has proven to be a suitable marker for measuring the n3 PUFA biostatus, which – for the reasons mentioned – also shows the lowest intra-individual variability compared to other markers such as plasma or plasma phospholipids. O3I levels can be categorized as desirable (≥8%), moderate (>6 to 8%), low (>4 to 6%), or very low (≤4%) ^(13)^. A target O3I value for a reduced risk for fatal coronary heart disease of ≥ 8% has been proposed ^(12)^, whereas individuals with O3I levels of ≤ 4% are at highest risk ^(12)^.

Despite its importance, few national health surveys have quantified n3 PUFA levels, mainly in blood plasma or phospholipids. The National Health and Nutrition Examination Survey (NHANES) reported plasma fatty acid levels as concentrations (µmol/L), whereas the Canadian Health Measures Survey used RBCs and reported national O3I levels ^(14)^. The latter found that the mean O3I was comparatively low at 4.5%. Data from other studies show that the average O3I is also in the low range in countries such as the USA, Italy, or Germany and not as high as in Japan or South Korea, where mean O3I levels are in a desirable range. Comprehensive data on the O3I status are not available in the United Kingdom (UK), however, with data from the UK Biobank (UKBB) now available ^(15,16)^, the n3 PUFA biostatus in that country may now theoretically be determined. However, the plasma FA data from the UKBB were expressed as concentrations or as a percent of total plasma fatty acids, so how these metrics compare with the O3I is unclear.

The first aim of our study was to develop an equation to estimate the O3I (eO3I) from the nuclear magnetic resonance (NMR) data of the UK Biobank so as to be able to compare UK O3I levels with those in other countries. The second aim was to examine the cross-sectional relations of the eO3I with important demographic (age, ethnic group, sex), anthropometric [body mass index (BMI), waist circumference (WC)] and lifestyle [fish oil supplement use, (oily) fish consumption, alcohol use, smoking, etc.] factors to help define the determinants of the eO3I. To accomplish these aims, we first performed an interlaboratory experiment to compare NMR-derived FA data with the O3I using gas chromatography (GC)-derived data. With a conversion equation thus generated, eO3I values were computed and then correlated in multivariable adjusted models with demographic and lifestyle variables.

## Methods

### UK Biobank

UK Biobank is a prospective, population-based cohort of approximately 500,000 individuals recruited between 2007 and 2010 at assessment centers across England, Wales and Scotland. Baseline data derived from questionnaires, biological samples and physical measurements were collected on all participating individuals, with longitudinal monitoring occurring via a mix of in-person and Electronic Medical Record data ^(15,16)^. The participants completed a touchscreen questionnaire, which collected information on socio-demographic characteristics, diet, and lifestyle factors. Anthropometric measurements were taken using standardized procedures. The touchscreen questionnaire and other resources are shown on the UK Biobank website (http://www.ukbiobank.ac.uk). UK Biobank has ethical approval (Ref. 11/NW/0382) from the North West Multi-centre Research Ethics Committee as a Research Tissue Bank (RTB). This approval means that researchers do not require separate ethical clearance and can operate under the RTB approval. All participants gave electronic signed informed consent. The UK Biobank study was conducted according to the guidelines laid down in the Declaration of Helsinki.

A random sample of approximately 125,000 participants was selected for biomarker assessment using nuclear magnetic resonance (NMR, Nightingale Health Plc, Helsinki, Finland)^(17)^ which included some plasma FAs. Usable NMR data were available from 117,938 participants. After removing individuals with missing information on BMI, socioeconomic status (SES) or alcohol use (n=830), the final sample size for this study was 117,108 (**Figure 1**).

**Figure 1:**
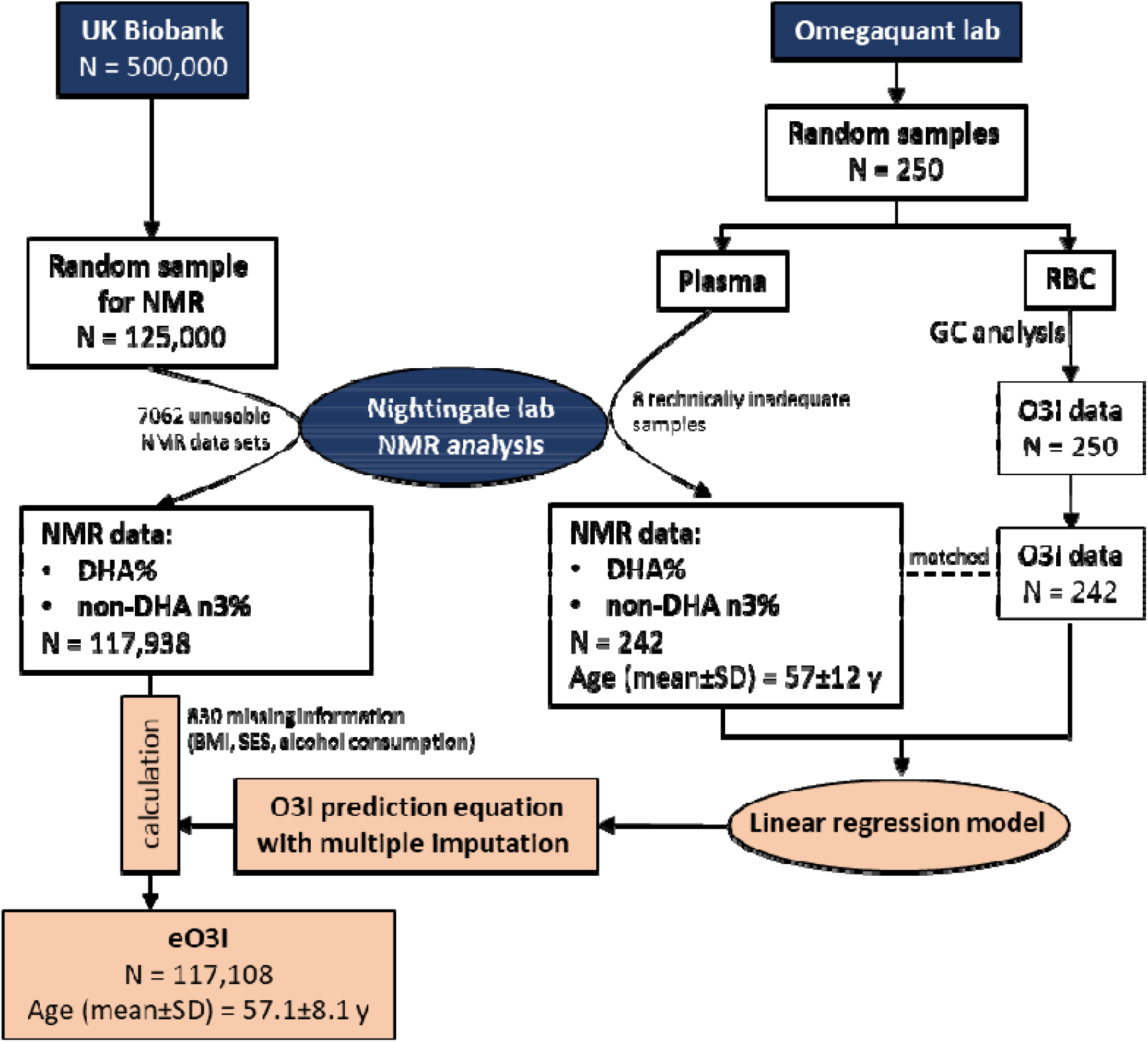
Flow chart for the interlaboratory experiment to create an Omega-3 Index (O3I) prediction equation and calculation of the estimated Omega-3 Index (eO3I) from the UK Biobank data (for details see text).

### Interlaboratory experiment

An interlaboratory experiment was undertaken to gather data for the creation of an O3I prediction equation (Figure 1). Random blood samples (n=250) received at OmegaQuant Analytics (OQA, Sioux Falls, SD, USA) for routine testing of the O3I were used for this study. EDTA blood tubes were spun to isolate the RBC fraction which was then analyzed for the O3I by GC as previously described ^(18)^. The plasma from these samples, which is normally discarded, was stored at -80C°. Once 250 had been collected (over about 2 weeks), the plasma aliquots were sent on dry ice for NMR analysis at the Nightingale lab. The data received from this analysis (n=242, 8 being technically inadequate) included multiple biometrics ^(19)^ which were subsequently used as predictors of the O3I as described below. The University of South Dakota Institutional Review Board reviewed this project and determined that because the project involved only deidentified samples and data, it did not meet the definition of “human subjects research” found in the federal regulations, and therefore did not require IRB approval.

### Statistical methods

#### Generating a prediction model for the estimated Omega-3 Index

As noted, the O3I is the sum of DHA and EPA in RBCs expressed as a percent of total RBC FAs. The relevant NMR data for this study were plasma DHA and total n3 PUFAs (each expressed as a percent of total FAs). No data on other n3 PUFAs were available. Thus, we began by examining predictions of the O3I by total n3 PUFA%, DHA% and non-DHA n3 PUFA% (i.e., total n3 PUFAs - DHA%). Quadratic terms for each predictor plus an interaction term were evaluated for evidence of improved fit using a significance level of 0.05. The remaining 246 biometric measurements ^(19)^ were then added to the model one at a time for evidence of improved fit, using a Bonferroni adjusted significance level of 0.05/246 = 0.0002 as the criterion for addition to the predictive model. Three individuals with extreme DHA%/non-DHA% were temporarily excluded from the analyses to improve model fitting (two individuals with DHA%<1 and one individual with DHA%>4 and non-DHA%>5) yielding a model building sample size of 239. The final model was evaluated using R-squared, Residual Standard Error and the distribution of residuals. Values of the eO3I were then imputed by making stochastic draws from the predicted sampling distributions (final predictive model) as described by Rubin for data missing at random ^(20)^. Sensitivity analyses were conducted considering extreme values of the observed DHA% and logarithmic transformations of the predictor and response variables.

### The eO3I and participant characteristics

We used eO3I data generated as described above to investigate its relationship with 14 sample characteristics: demographic (sex, age, ethnic group, deprivation index, urbanicity, education), anthropometric (BMI, WC), dietary (frequency of oily and non-oily fish consumption and regular fish oil use), and behavioral (smoking, alcohol use, exercise) (**Table 1**). BMI was classified according to the World Health Organization (WHO) into “normal weight”, “overweight” and “obesity”. For WC categories, the cohort was divided into quartiles. Information on fish portion sizes and fish oil supplement EPA+DHA content was not collected. Townsend deprivation index scores were derived from national census data about car ownership, household overcrowding, owner occupation, and unemployment aggregated for residential postcodes ^(21)^. Higher deprivation index scores indicate greater degrees of socioeconomic deprivation. Our analysis used national quintiles of the deprivation index instead of continuous scores ^(22)^. Four groups of education were formed: College (or University degree, other professional qualifications), Associates (A or AS level or equivalent), SEs (O level, General Certificate of Secondary Education, Certificate of Secondary Education, National Vocational Qualifications, Higher National Diploma, Higher National Certificate or equivalent), none. We analyzed the bivariate relationships between the eO3I and each of the 14 sample characteristics considering multiple imputations in the estimation of standard errors. A fully adjusted linear model predicted the eO3I by each of the 14 sample characteristics simultaneously, again accounting for multiple imputation in the estimation of standard errors. Model *R*^*2*^ was computed. In order to estimate the contribution of each predictor towards model *R*^*2*^, each of the 14 sample characteristics was then removed from the model (one at a time) and computing the difference in *R*^*2*^ between the full and drop-one model. Parallel analyses were conducted for plasma percent DHA (direct NMR measurement), without the need for multiple imputation. All analyses were conducted in R version 4.1.2. and used a 0.05 statistical significance threshold.

**Table 1:**
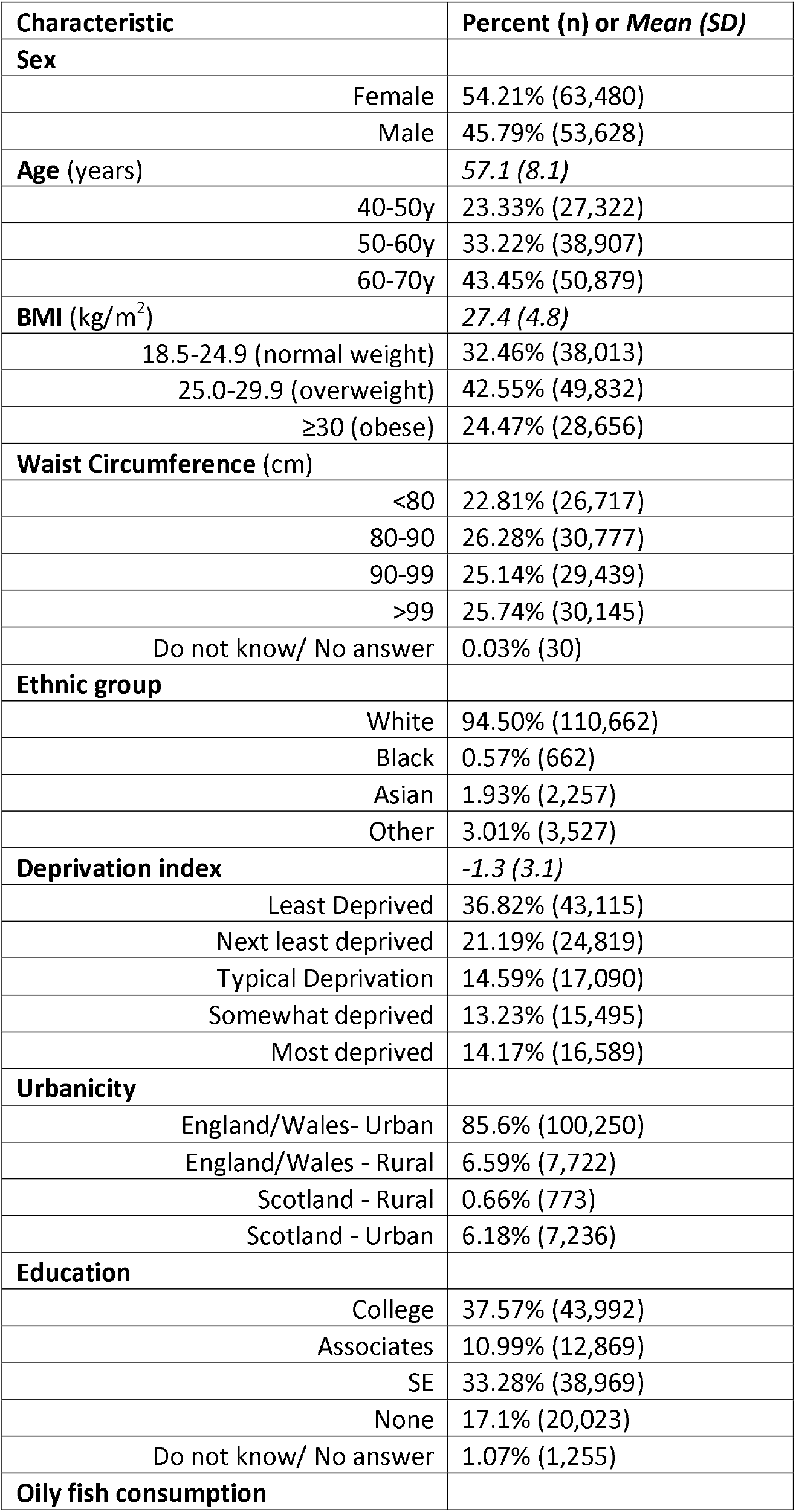
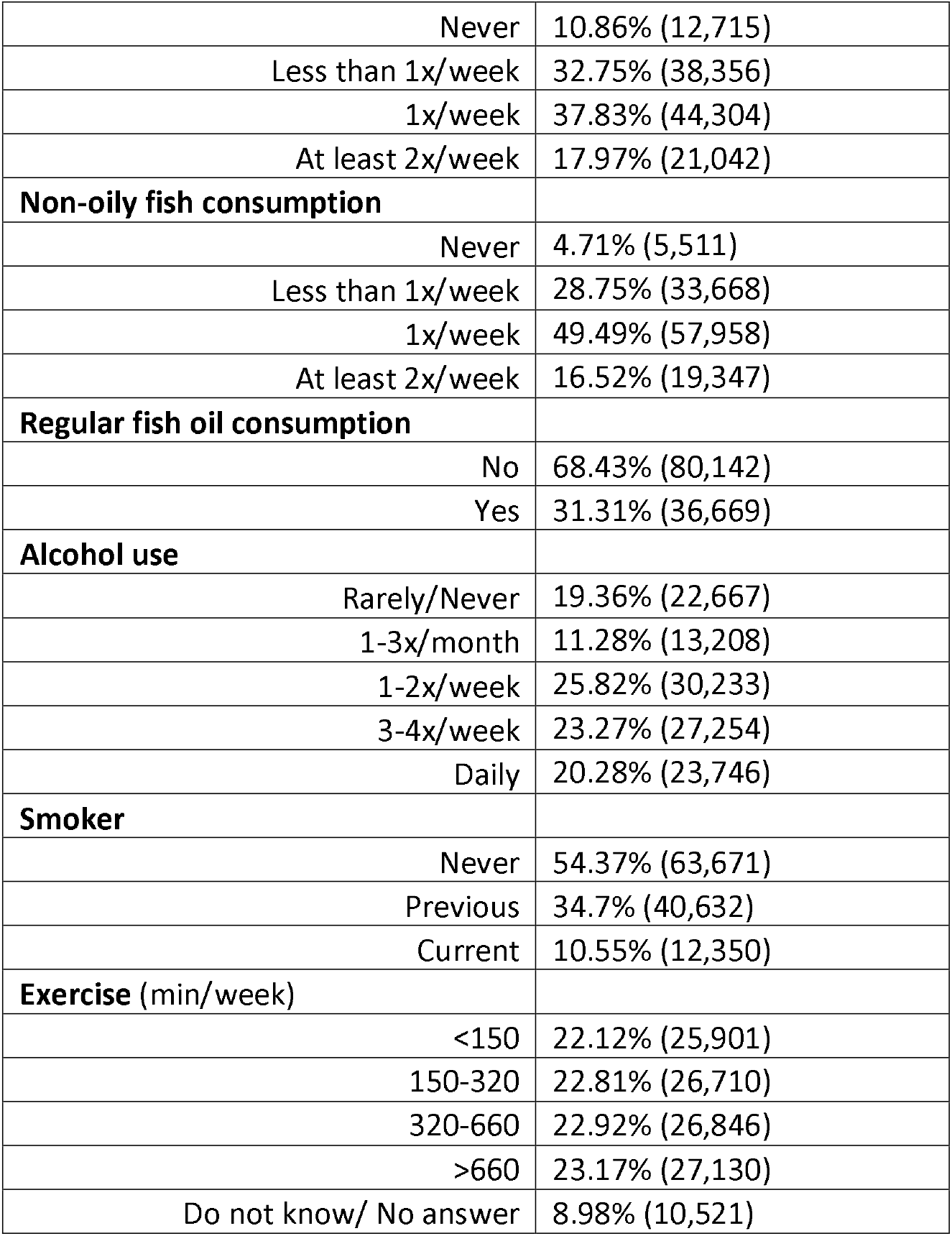
Demographics of the UK Biobank sample (n=117,108).

## Results

### Generating a prediction equation based on the interlaboratory experiment

In models predicting the O3I by a single variable, percent DHA was a better predictor (*r*^*2*^=0.65), than total n3 (*r*^*2*^=0.60). We then added non-DHA% to the model with DHA%, yielding a model with *r*^*2*^=0.67. Residuals showed a reasonably normal distribution (**supplemental Figure 1**). Quadratic terms and an interaction between DHA% and non-DHA% were added to the model, but none showed evidence of improvement [p=0.41 (quadratic DHA%); p=0.80 (quadratic non-DHA%); p=0.20 (interaction DHA and non-DHA%)]. Log transforming the O3I and/or DHA% and non-DHA% prior to fitting the regression equation did not improve the *r*^*2*^ values (61-65%) with little visible improvement in the normality of outliers (details not shown). Thus, the untransformed, additive model with two terms was determined to be the final prediction model (**Table 2**). The correlation of the eO3I vs observed O3I values was 0.823 on the complete dataset of n=242, including three extreme values which were not included in the initial model-building step (**Figure 2**).

**Table 2:** Regression model predicting the Omega-3 Index (n=239).

| Predictor | Beta (SE) | P-value |
| --- | --- | --- |
| Intercept | -0.1014 (0.315) | 0.75 |
| NMR DHA% | 2.6290 (0.194) | $<2 \times 10^{-16}$ |
| NMR non-DHA% | 0.4673 (0.126) | 0.00027 |
| Model summary statistics: Residual SE (1.217); $r^2=0.673$; $r=0.823$ | | |

**Figure 2:**
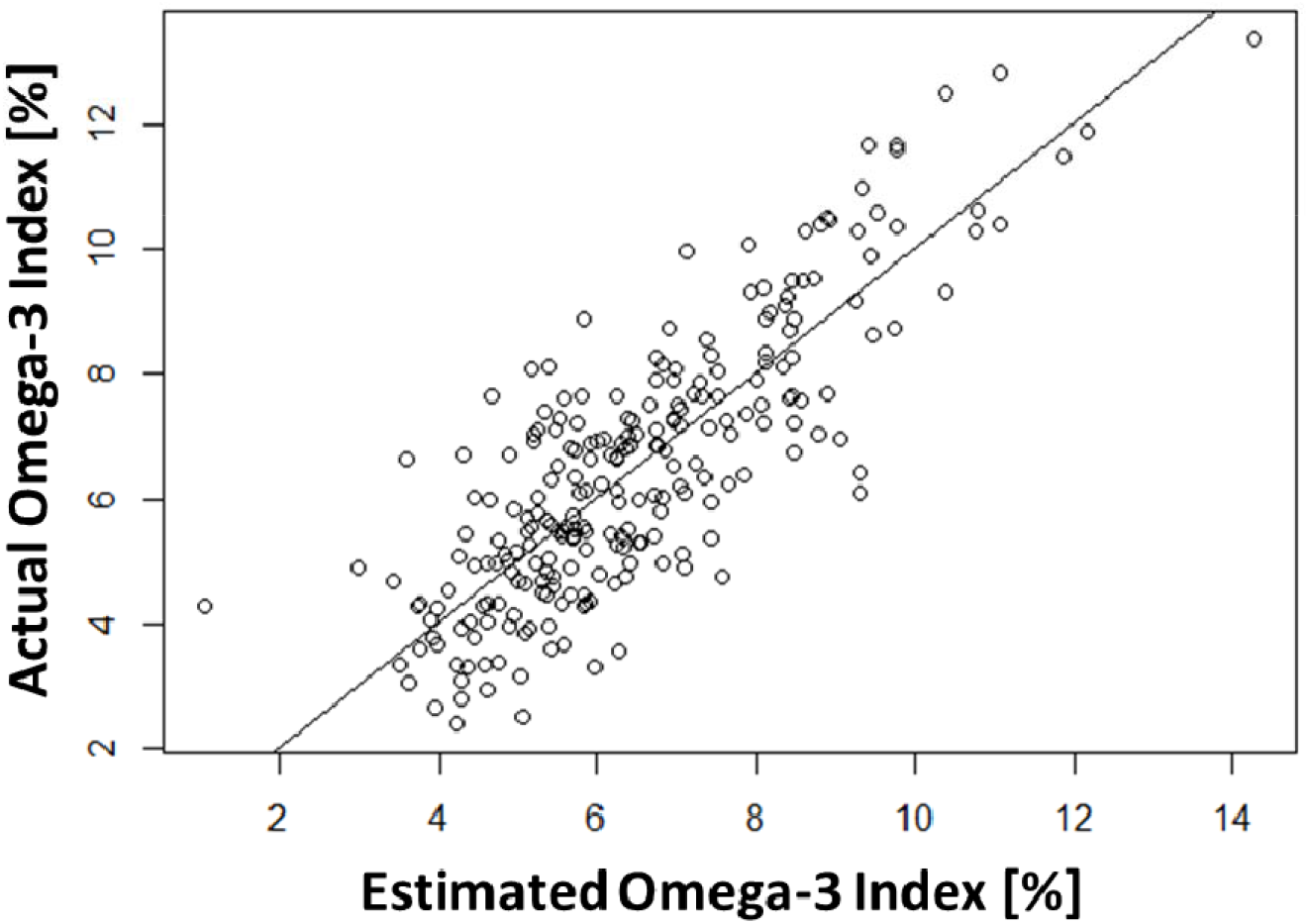
Predicted Omega-3 Index (eO3I) vs actual Omega-3 Index (O3I). The line of identity (y=x) is plotted. All values are percent of total RBC FAs. Final *r*=0.823.

### Demographics of the UK Biobank sample

The demographics of the analyzed UK Biobank sample are in Table 1. The cohort had a mean age of 57 years with slightly more women than men. The individuals were overwhelmingly white and lived in urban areas. The mean deprivation index was -1.3, meaning that, on average, this sample was somewhat less deprived than typical in the UK (which would be an index of 0). The plurality of individuals were overweight, followed by normal weight and then obese.

Most individuals in the cohort stated that they regularly ate fish. Non-oily fish is slightly preferred compared to oily fish. Almost half of the individuals stated that they eat non-oily fish 1x/week. However, the proportion of individuals who eat oily fish 1x/week is also quite high at 38%. Regular fish oil supplement use was reported by 31%. Most of the study participants reported some regular alcohol use. More than half of the individuals were non-smokers, some were ex-smokers and a few continued to smoke.

### Predicted Omega-3 Index in the UK Biobank sample

In the interlaboratory experiment, the mean (SD) of plasma DHA% from the NMR analysis was 2.1% (0.6%) and the plasma non-DHA% was 2.1% (0.9%). The mean (SD) GC-based O3I [6.5% (2.2%)] was (naturally) the same as that estimated from the regression equation from the NMR data. All three measurements showed modest right-skewness.

The mean eO3I was 5.58% for all 117,938 individuals in this UKBB cohort. We used multiple (i.e., 10) imputations for each person using the prediction equation in Table 2 in order to compute the SDs. The SD ignoring the multiple imputation step was 1.89%, whereas with imputations it was 2.25%, reflecting the variability from different analytical methods (NMR vs GC), different sample types (plasma vs RBC), and different n3 PUFAs (DHA vs EPA+DHA).

### Correlates of the estimated Omega-3 Index in the UK Biobank sample (Table 3)

With the exceptions of rurality/urbanicity, virtually every metric included in **Table 3** was statistically significantly correlated with the eO3I (p<0.0001). For the *behavior-independent* demographic variables, both **sex** and **age** were strongly associated with the eOI3 (Table 3 and **Figure 3a**). The eO3I in men was -0.62 percentage points lower than in women before, and - 0.38% lower after adjustment. Additionally, the eO3I increased with age. Compared to the 40-50 year-age-group (Reference group), the eO3I in the 50-60 year-, and 60-70 year-age-group increased by 0.29% and 0.64% respectively. After adjustment, the eO3I was still directly associated with age (0.14% and 0.34%, respectively).

**Table 3:** Associations of demographic characteristics and the estimated Omega-3 Index (eO3I) in unadjusted and adjusted (for all variables in Table 1) analyses (n=117,108).

| Characteristics | eO3I<br>Mean<br>(SD) | eO3I<br>Unadj Diff or Corr<br>(95% CI) | Unadj P-value | eO3I<br>Adj Diff or Corr<br>(95% CI) | Adj P-value |
| --- | --- | --- | --- | --- | --- |
| **Sex** |  |  |  |  |  |
| Female | 5.9 (2.2) | 0 |  | 0 |  |
| Male | 5.2 (2.2) | -0.62 (-0.65,-0.59) | <0.0001*** | -0.38 (-0.42,-0.34) | <0.0001*** |
| **Age (years)** |  |  |  |  |  |
| 40-50 (reference) | 5.2 (2.1) | 0 |  | 0 |  |
| 50-60 | 5.5 (2.2) | 0.29 (0.25,0.33) | <0.0001*** | 0.15 (0.11,0.19) | <0.0001*** |
| 60-70 | 5.8 (2.3) | 0.64 (0.6,0.68) | <0.0001*** | 0.35 (0.31,0.39) | <0.0001*** |
| **BMI (kg/m<sup>2</sup>)** |  |  |  |  |  |
| 18.5-24.9 (normal weight; reference) | 6 (2.3) | 0 |  | 0 |  |
| 25.0-29.9 (overweight) | 5.6 (2.2) | -0.4 (-0.43,-0.36) | <0.0001*** | -0.13 (-0.18,-0.09) | <0.0001*** |
| ≥30 (obese) | 5.1 (2.1) | -0.88 (-0.92,-0.84) | <0.0001*** | -0.38 (-0.44,-0.32) | <0.0001*** |
| **Waist Circumference (cm)** |  |  |  |  |  |
| <80 (reference) | 6.2 (2.3) | 0 |  | 0 |  |
| 80-90 | 5.8 (2.2) | -0.38 (-0.42,-0.33) | <0.0001*** | -0.22 (-0.27,-0.17) | <0.0001*** |
| 90-99 | 5.4 (2.2) | -0.72 (-0.77,-0.67) | <0.0001*** | -0.37 (-0.43,-0.31) | <0.0001*** |
| >99 | 5.0 (2.1) | -1.13 (-1.18,-1.08) | <0.0001*** | -0.55 (-0.63,-0.47) | <0.0001*** |
| Do not know/No answer | 4.9 (1.7) | -1.23 (-2.24,-0.21) | 0.018* | -0.8 (-1.78,0.17) | 0.1 |
| **Ethnic group** |  |  |  |  |  |
| White (reference) | 5.6 (2.2) | 0 |  | 0 |  |
| Black | 5.7 (2.3) | 0.09 (-0.12,0.31) | 0.41 | 0.17 (-0.04,0.38) | 0.12 |
| Asian | 5 (2.3) | -0.58 (-0.7,-0.46) | <0.0001*** | 0 (-0.12,0.12) | 0.97 |
| Other | 6.5 (2.6) | 0.94 (0.85,1.04) | <0.0001*** | 1.03 (0.93,1.12) | <0.0001*** |
| **Deprivation index** |  |  |  |  |  |
| Least Deprived (reference) | 5.7 (2.2) | 0 |  | 0 |  |
| Next least deprived | 5.6 (2.2) | -0.09 (-0.13,-0.04) | 0.0001** | -0.01 (-0.06,0.03) | 0.49 |
| Typical Deprivation | 5.5 (2.2) | -0.2 (-0.25,-0.15) | <0.0001*** | -0.07 (-0.12,-0.02) | 0.003** |
| Somewhat deprived | 5.4 (2.2) | -0.33 (-0.38,-0.28) | <0.0001*** | -0.12 (-0.17,-0.07) | <0.0001*** |
| Most deprived | 5.3 (2.3) | -0.42 (-0.48,-0.37) | <0.0001*** | -0.12 (-0.18,-0.07) | <0.0001*** |
| **Urbanicity** |  |  |  |  |  |
| England/Wales- Urban (reference) | 5.6 (2.2) | 0 |  | 0 |  |
| England/Wales – Rural | 5.8 (2.2) | 0.22 (0.16,0.28) | <0.0001*** | 0.02 (-0.04,0.08) | 0.5 |
| Scotland – Rural | 5.6 (2.3) | 0.01 (-0.19,0.2) | 0.96 | 0.03 (-0.15,0.21) | 0.74 |
| Scotland – Urban | 5.5 (2.2) | -0.1 (-0.16,-0.03) | 0.0027** | -0.06 (-0.12,0) | 0.049* |
| **Education** |  |  |  |  |  |
| College (reference) | 5.8 (2.3) | 0 |  | 0 |  |
| Associates | 5.7 (2.2) | -0.14 (-0.2,-0.09) | <0.0001*** | -0.08 (-0.13,-0.03) | 0.00094** |
| SEs | 5.4 (2.2) | -0.37 (-0.41,-0.34) | <0.0001*** | -0.18 (-0.22,-0.15) | <0.0001*** |
| None | 5.4 (2.2) | -0.42 (-0.47,-0.38) | <0.0001*** | -0.23 (-0.27,-0.18) | <0.0001*** |
| Do not know/No answer | 5.4 (2.2) | -0.39 (-0.54,-0.23) | <0.0001*** | -0.26 (-0.4,-0.11) | 0.00056** |
| **Oily fish consumption** |  |  |  |  |  |
| Never (reference) | 4.3 (1.7) | 0 |  | 0 |  |
| Less than 1x/week | 5 (1.9) | 0.77 (0.71,0.82) | <0.0001*** | 0.47 (0.41,0.53) | <0.0001*** |
| 1x/week | 5.8 (2.1) | 1.56 (1.51,1.62) | <0.0001*** | 1.11 (1.05,1.17) | <0.0001*** |
| At least 2x/week | 6.9 (2.6) | 2.64 (2.57,2.7) | <0.0001*** | 2.12 (2.05,2.18) | <0.0001*** |
| **Non-oily fish consumption** |  |  |  |  |  |
| Never (reference) | 4.3 (2) | 0 |  | 0 |  |
| Less than 1x/week | 5.3 (2.2) | 1 (0.92,1.07) | <0.0001*** | 0.34 (0.26,0.42) | <0.0001*** |
| 1x/week | 5.7 (2.2) | 1.38 (1.31,1.46) | <0.0001*** | 0.35 (0.26,0.43) | <0.0001*** |
| At least 2x/week | 6.1 (2.4) | 1.79 (1.71,1.87) | <0.0001*** | 0.49 (0.4,0.58) | <0.0001*** |
| **Regular Fish oil consumption** |  |  |  |  |  |
| No (reference) | 5.3 (2.1) | 0 |  | 0 |  |
| Yes | 6.2 (2.3) | 0.97 (0.93,1) | <0.0001*** | 0.68 (0.65,0.71) | <0.0001*** |
| **Alcohol use** |  |  |  |  |  |
| Rarely/Never (reference) | 5.4 (2.3) | 0 |  | 0 |  |
| 1-3x/month | 5.4 (2.2) | -0.02 (-0.08,0.04) | 0.61 | 0.01 (-0.05,0.07) | 0.69 |
| 1-2x/week | 5.5 (2.2) | 0.14 (0.09,0.19) | <0.0001*** | 0.11 (0.06,0.16) | <0.0001*** |
| 3-4x/week | 5.8 (2.2) | 0.38 (0.33,0.43) | <0.0001*** | 0.26 (0.21,0.31) | <0.0001*** |
| Daily | 5.8 (2.2) | 0.4 (0.35,0.45) | <0.0001*** | 0.3 (0.25,0.35) | <0.0001*** |
| **Smoker** |  |  |  |  |  |
| Never (reference) | 5.7 (2.3) | 0 |  | 0 |  |
| Previous | 5.6 (2.3) | -0.04 (-0.08,-0.01) | 0.025* | -0.03 (-0.07,-0) | 0.07 |
| Current | 4.8 (2) | -0.86 (-0.92,-0.8) | <0.0001*** | -0.58 (-0.64,-0.53) | <0.0001*** |
| **Exercise (min/week)** |  |  |  |  |  |
| <150 (reference) | 5.5 (2.2) | 0 |  | 0 |  |
| 150-320 | 5.7 (2.2) | 0.23 (0.18,0.27) | <0.0001*** | 0.01 (-0.03,0.05) | 0.68 |
| 320-660 | 5.7 (2.3) | 0.26 (0.21,0.3) | <0.0001*** | -0.04 (-0.08,0.01) | 0.092 |
| >660 | 5.6 (2.3) | 0.12 (0.07,0.16) | <0.0001*** | -0.16 (-0.21,-0.12) | <0.0001*** |
| Do not know/No answer | 5.3 (2.2) | -0.17 (-0.17,-0.04) | 0.00086** | -0.1 (-0.16,-0.04) | 0.00074** |

**Figure 3:**
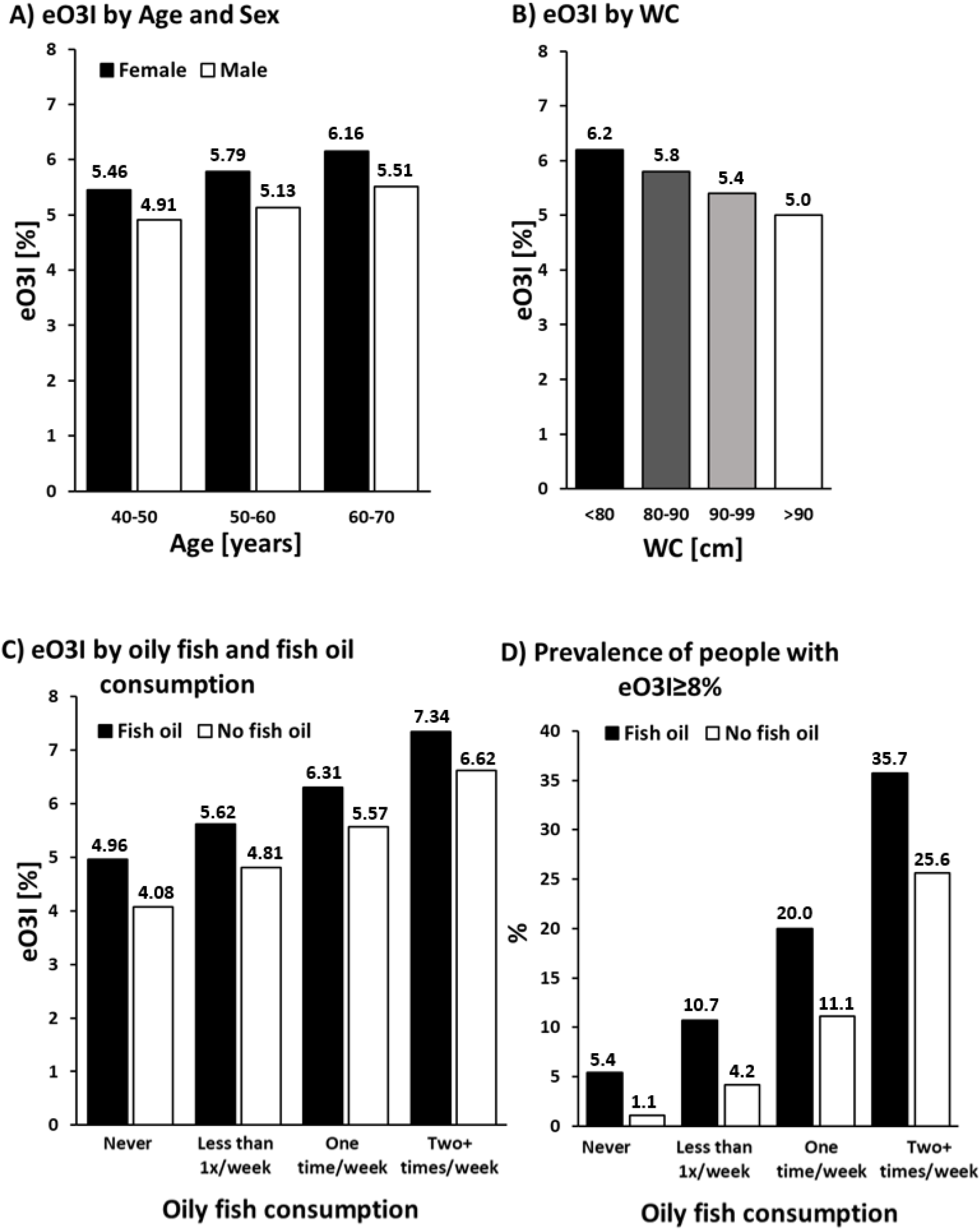
Predicted Omega-3 Index (eO3I) by A) age and sex, B) waist circumference (WC), C) oily fish and fish oil consumption. D) Percentage of people with an eO3I≥8% depending on oily fish and fish oil consumption. All pairwise comparisons (male vs. female in each age group, sex-specific age groups, all WC categories, fish oil vs. no fish oil in different oily fish consumption groups and between different oily fish consumption groups) were p<0.0001.

There was an inverse association between the eO3I and **BMI**, both before and after adjustment (Table 3). Compared to normal weight people, the mean eO3I was 0.4% and 0.88% lower in the overweight and obese individuals, respectively. These differences were somewhat attenuated after adjustment but remained -0.13% or -0.38%, respectively.

Associations between **WC** and eO3I were examined as a further body fat variable (Table 3 and Figure 3b). Compared to the reference group, there was a monotonic decrease in the eO3I across WC quartiles. In the group with the highest WC, the eO3I was reduced by 1.13% (adjusted, -0.55%) compared to the reference group.

With regards to **ethnic group**, the eO3I in Blacks and Asians was not significantly different from that in Whites. “Other” ethnic groups (3% of the total) had higher eO3I than Whites (0.94%, adjusted, 1.03%).

SES (as reflected by the **deprivation index**) was inversely associated with the eO3I. Compared to the group of least deprived individuals, the eO3I decreased continuously as the **deprivation index** increased. The most deprived group showed a 0.13% lower eO3I after adjustment compared to the least deprived group.

Another SES-related metric is the level of **education**. Compared to the group with the highest professional qualification (College, reference), the eO3I was lower in each successive group of educational achievement. At the extremes, individuals with the least education had a mean eO3I of -0.42% (adjusted, -0.23%) lower than the reference group.

As expected, regular consumption of **oily fish** and taking **fish oil supplements** were the strongest *behaviour-related* associations with the eO3I (Table 3, Figure 3c). The mean eO3I of people who take fish oil and eat oily fish at least 2x/week (n=8279) was 7.34%, but only 36% of them had an eO3I ≥8% (Figure 3d). The 14 characteristics examined in this study together explained about 21% of the variability in the eO3I (**Table 4**). Regular oily fish intake alone explained 7%, or about 34% of the total. Compared to non-fish eaters (reference), even less than one serving of oily fish per week was associated with a 0.77% higher eO3I (0.47% after adjustment). As oily fish consumption continued to increase, so did the eO3I, with 1 portion of oily fish per week being associated with a 1.56% higher eO3I (adjusted, 1.11%), and 2.64% higher eO3I (adjusted, 2.12%) with 2 or more portions of oily fish per week. **Non-oily fish consumption** was more weakly associated with the eO3I, explaining less than 1% of the eO3I variability (Table 4). After adjustment, the difference in eO3I between non-fish eaters and those consuming at least 2 portions of non-oily fish per week was only 0.49%.

**Table 4:** R-squared values for the participant’s characteristics on the variability of eO3I.

|  |  |  |
| --- | --- | --- |
| **Overall model** | 20.5% |  |
| **Variable dropped<sup>1</sup>** | **New R-squared<sup>1</sup>** | **Uniquely explained variability<sup>2</sup>** |
| **Oily fish consumption** | 13.5% | 7.0% |
| **Regular fish oil consumption** | 18.7% | 1.8% |
| **Smoking** | 19.9% | 0.6% |
| **Ethnic group** | 19.9% | 0.6% |
| **Sex** | 20.1% | 0.4% |
| **Age** | 20.2% | 0.3% |
| **Alcohol use** | 20.2% | 0.3% |
| **Waist Circumference** | 20.2% | 0.3% |
| **BMI** | 20.3% | 0.2% |
| **Education** | 20.3% | 0.2% |
| **Non-oily fish consumption** | 20.4% | 0.1% |
| **Exercise** | 20.4% | 0.1% |
| **Deprivation index** | 20.5% | 0.0% |
| **Urbanicity** | 20.5% | 0.0% |

1. The new model R-squared for the remaining 13 variables when predicting eO3I. Variables are ranked according to their independent contribution on the total variability.
2. Uniquely explained variability is computed as the difference in the model without the variable compared to the full model with all 14 variables (overall model).

On average, individuals who reported regular **fish oil supplement** use had a 0.97% higher eO3I than individuals who didn’t. Even after adjustment for all the other factors in Table 3, it was still 0.68% higher. The impact of fish oil supplementation on the eO3I was additive with the number of oily fish servings per week (Figure 3c). People who reported taking a fish oil supplement and consuming two or more oily fish meals per week had a mean eO3I of 7.3% (2.7%) compared with 4.1% (1.7%) for those at the other extreme consuming neither (p<0.0001). Fish oil intake explained ∼9% of the total explained variability in the eO3I (Table 4).

**Alcohol** use also showed associations with the n3 PUFA biostatus. Compared to the group with the lowest alcohol use (none, reference group), the eO3I was increased with higher alcohol use. The highest alcohol use (daily) had a 0.4% higher eO3I (adjusted, 0.3%) compared to the reference group.

**Smoking** was negatively associated with the eO3I. Smokers had a 0.86% lower eO3I compared to non-smokers, even after adjustment (−0.58%).

The level of **exercise** showed a weak but still significant inverse relationship to the eO3I. Compared to the group with the lowest level of physical activity (reference group), the eO3I was only lower in the group with the highest level of physical activity (adjusted -0.15%).

The plasma DHA% measured by NMR showed the same patterns/relationships with regards to all 14 characteristics as did the eO3I (**Supplemental Table 1**). However, all together, these predictors accounted for approximately 30% of the variability in DHA% (**Supplemental Table 2**), compared to only around 20% for the eO3I (Table 4). For both DHA and eO3I, oily fish consumption and fish oil supplement use explained most of the variation.

## Discussion

In order to be able to investigate relationships between n3 PUFAs and health or disease measures on populational level, it is preferable to use a biomarker of n3 PUFA status rather than an estimate of dietary intake from a questionnaire. This is because biomarker levels are objectively measured, precise, and reflect not only dietary intake but *in vivo* metabolic conversions that cannot be captured with memory-based dietary intake surveys. The UK Biobank provides a large database to investigate such associations due to its extensive health data and number of participants. Since the O3I, which is a common metric to evaluate the n3 PUFA biostatus, was not measured directly in the UK Biobank, our goal was to develop a prediction model for the O3I in order to convert the data that do exist on n3 PUFA biostatus (plasma DHA% and total n3% from NMR) into the eO3I. We found very good agreement (r=0.82) between the estimated O3I values (eO3I) and the actual O3I. The eO3I equation was built on data from 242 samples, which were not included in the UK Biobank. We assumed that since the same lab (Nightingale) and method (NMR) were used for the UK Biobank analyses and for our interlaboratory test that the conversion equation should be applicable to UK Biobank data.

### eO3I in the UK Biobank in comparison with other countries

In the UK there are no datasets from which a national, average O3I can be determined. A global survey on n3 PUFA levels in different countries reported “very low” n3 PUFA levels in RBCs (<4% EPA + DHA of total FAs) for the UK using extrapolated data from 5 studies including 461 individuals ^(23)^. The mean eO3I in the UK Biobank cohort of 117,938 individuals was 5.58%, and thus, significantly higher than that estimated by Stark et al. The eO3I in the UK Biobank is comparable to that of countries such as Germany (5.8%) or the USA (5.44%), as shown in a recent report ^(13)^. Nevertheless, an average O3I of 5.58% is still well below the optimal of ≥8%. People who regularly eat oily fish and supplement fish oil have a significantly better n3 PUFA biostatus. The mean eO3I of people who take fish oil and eat oily fish at least 2x/week (n=8279) was 7.34% (2.66%), and 35.7% (n=2957) of them had an eO3I ≥8%. Thus, even among this group with the highest oily fish + supplement use, an optimal eO3I was not the norm.

### Predictors of the eO3I in the UK Biobank cohort

Various factors besides EPA+DHA intake can affect O3I levels. Several other authors have explored this question ^(24–27)^. Since not all comparative studies in the literature used the O3I as a metric, the general term “n3 PUFA biostatus” is used below, which means the eO3I in relation to this study.

A higher n3 PUFA biostatus in **men compared to women** has been observed previously ^(28,29)^. It has been suggested that a healthier diet (e.g., higher consumption of fish or plant n3 PUFAs) and lifestyle (e.g., no smoking) could be a reason for the higher n3 PUFA biostatus in women ^(28)^. However, the influence of factors such as smoking or (oily) fish and fish oil supplement intake and obesity were all considered in our model, and even after the adjustment there remained a significant and biologically relevant difference in the eO3I between the sexes. Other factors independent of lifestyle (e.g., genetics and metabolism) may also play a role. The reason for a higher n3 PUFA biostatus in females could be due to enhanced production of EPA + DHA from the precursor FA alpha-linolenic acid (ALA, C18:3), a conversion mediated at least in part by estrogen ^(30–32)^. In a series of studies, Burdge and Wootton ^(31,33)^ concluded that estrogen up-regulates delta-6 desaturase, which is the rate-limiting step in the conversion from ALA to EPA and DHA. However, dietary intakes of n3 PUFAs were not controlled for over the entire study period, whereby a dietetic effect on the intake of preformed EPA+DHA cannot be ruled out ^(34)^. In a study by Giltay et al. ^(30–32)^ in postmenopausal women who received hormone replacement therapy, the level of DHA in plasma cholesteryl esters increased by 20%, which was attributed to the estrogenic effect.

Although not addressed in this study, **genotype** could play a role in determining n3 PUFA biostatus. This, however, is controversial with some studies reporting an effect of a “heritability” score ^(28)^, on the O3I whereas others found no association with the O3I of any given single nucleotide polymorphism ^(35)^. We hope to investigate a possible genetic influence on the level of eO3I in future UK Biobank studies.

An **age-related influence** on the n3 PUFA biostatus is also known and has been established in several studies ^(27,28,36)^. The reason for increased n3 PUFA biostatus in older individuals may be a function of decreased n3 PUFA turnover in tissues in older individuals ^(37)^, but it does not appear to simply be the result of higher fish intake or supplement use as these were included in the adjusted model.

Our data are in line with comparable epidemiological studies which also found inverse associations between **anthropometric markers** such as the **BMI** ^(38)^ or **WC** ^(28,36)^ and n3 PUFA biostatus. In obese individuals, the eO3I in the present cohort was 0.38% lower than that of normal weight individuals. The associations between WC and n3 PUFA biostatus appeared to be even stronger than those with BMI with a 0.55% lower eO3I in the group with highest WC compared to the reference group. Since WC is a direct reflection specifically of abdominal obesity, WC is now thought to be superior to BMI as a predictor of cardio-metabolic diseases ^(39)^. As to potential mechanisms, Cazolla et al. proposed that increased oxidative stress in obese individuals could lead to a reduction in the levels of EPA and DHA in RBC membranes ^(40)^. This was based on their observation that RBCs from obese individuals were more susceptible to oxidative stress than those from normal weight subjects. A disturbed hepatic PUFA metabolism may also contribute to a lower n3 PUFA biostatus. Animal studies found that obesity and diabetes resulted in reduced expression of key enzymes involved in the synthesis of EPA+DHA from ALA ^(41)^.

Several lifestyle variables were also associated with n3 PUFA biostatus. Consistent with the literature ^(24,28,36,42–44)^, we found a strong negative influence of **smoking** on the n3 PUFA biostatus. Increased oxidative stress and lipid peroxidation, caused by smoking, may destroy long chain PUFAs such as EPA and DHA in cell membrane phospholipids and, thus, influence their levels in the body ^(45,46)^. The possibility that this relationship is explained by differences in fish consumption and/or supplement use between smokers and non-smokers seems unlikely given the inclusion of these variables in the adjusted analysis. We also observed that the n3 PUFA biostatus was directly related to **alcohol** use, since the eO3I increased along with the reported frequency of alcohol use. Some studies also found alcohol use as an independent predictor of the n3 PUFA biostatus ^(47)^, while others did not ^(24,36)^. In the study by di Giuseppe, a positive influence of alcohol was only found in women and predominantly with wine drinking (and not beer or spirits). Since the UK Biobank does not provide information on the type of alcoholic beverages, a further differentiated analysis is not possible at this point. One can speculate that individuals that drink more alcohol may also eat more fish, and there was a modest association between oily fish eaters and drinking alcohol (e.g., 60% of daily alcohol users eat oily fish at least once/week vs 50% of monthly/hardly ever alcohol users). However, as with all of the potential determinants of the eO3I discussed here, significant associations with alcohol use remained after multivariable adjustment. In contrast, **exercise** in the UK Biobank cohort was only weakly associated with n3 PUFA biostatus. Small but significant differences were only found between the group with the highest and lowest reported minutes per week of physical exercise. In line with our results, a previous study ^(48)^ found significantly lower O3I levels in German national elite winter endurance athletes (mean O3I: 4.97%) or National Collegiate Athletic Association Division I athletes in the US (mean O3I: 4.33%) ^(49)^ compared to the general population in Germany (mean O3I: 5.8%) and the US (mean O3I: 5.44%) ^(13)^. In contrast to our results, a cross-sectional study showed that exercise time, exercise capacity and heart rate recovery strongly correlated with the O3I in patients with coronary artery disease ^(50)^.

The consumption of preformed **EPA and DHA** has the greatest influence on the n3 PUFA biostatus, as various other studies have already shown ^(24,28,36,38)^. The highest vs the lowest oily fish intake was associated with an unadjusted increase in the eO3I of 2.6%, decreasing only to 2.2% after adjustment. Fish oil supplement use had the second greatest effect in our study (i.e., 1% increase; 0.67% adjusted). The adjusted value was approximately equal to that (0.5%) of consuming <4 servings of oily fish per month. Since daily consumption of the most generic fish oil supplement would provide about 300 mg of EPA+DHA per day (about 2100 mg per week), and one serving of an oily fish like salmon would provide roughly 1000 mg of EPA+DHA per week, the smaller effect of on the eO3I of reported supplement use here suggests that the respondents in this cohort may have either overestimated what “regular” fish oil consumption means or underestimated their fish intake. These limitations are elaborated on below. Suffice it to say, that as noted above, people reporting both >2 oily fish meals per week AND fish oil supplement use had an adjusted eO3I of 7.34% compared to the 4.08% of the reference group consuming no fish or supplements.

The **deprivation index** – a measure of SES – was also identified as an inverse and independent predictor of the n3 PUFA biostatus confirming past studies (Harris et al. 2012). Individuals with higher SES have a better health behavior, a lower BMI, and smoke less ^(51)^, and they eat healthier food, which also includes higher fish and seafood consumption ^(52,53)^. However, several of these factors were considered in our adjusted model, and the inverse relationship between the eO3I and the deprivation index remained significant. We also identified **education** as a strong independent predictor for eO3I as have others (Wagner et al. 2015). The association between education and the eO3I was still significant even after adjustment for the deprivation index.

### Strength and limitations

The strength of the study is clearly the very large and fairly well-characterized cohort and an objectively measured biomarker of n3 PUFA status. However, the study has a number of potential limitations. First, the eO3I had to be extrapolated from plasma n3 PUFA measures done by NMR. The imprecision arising from this extrapolation (accounted for with multiple imputations of the eO3I) diminished the ability of the suite of variables we included to predict n3 PUFA biostatus. Specifically, the 14 factors explained 30% of the variability in plasma DHA% but only 20% of the eO3I variability. Thus, some relationships may have been missed as a result of using the eO3I. Another limitation is the relatively narrow age range of participants, from 40 to 70 years. Thus, our data on the eO3I cannot be extrapolated to other age groups. Obviously, these data only apply to the UK, or perhaps “western” populations in general (given the similarity in eO3I values seen here and in other countries ^(13)^; but they do not apply to countries with diets and lifestyles differing markedly from those in the UK.

Finally, self-reported fish consumption data is prone to under- and over-reporting. Given a general understanding of “fish as a healthy food”, people are more likely to over-report their fish consumption. In the UK Biobank study, no portion sizes of oily and non-oily fish consumption were collected which could, theoretically, have allowed more precise associations between fish intake and eO3I to be observed. In addition, no other data on fish oil supplement intake was collected (e.g., dosage, potency, and frequency of intake). These variables therefore only provided a rough estimate of the actual consumption of fish and fish oil. As usual, data from cross-sectional studies cannot reveal causal relationships. Our data are thus only hypothesis-generating.

### Conclusion

The results of this investigation allow for the first time an estimation of the average O3I in the UK. Secondly, they largely confirm previously observed determinants of the O3I, namely oily fish consumption, fish oil intake, sex, age, WC, BMI, and various lifestyle variables. A recent study with the UK Biobank showed that fish oil supplementation was associated with reduced risk for CVD outcomes and all-cause mortality ^(54)^. With the ability to derive an eO3I as a metric for the n3 PUFA biostatus, it will now be possible to investigate its relationship with risk for multiple diseases such as CVD, type 2 diabetes mellitus, or neurodegenerative disorders in the UK Biobank.

## Data Availability

All data produced in the present study are available upon reasonable request to the authors

## Authorship

JPS: Investigation, Conceptualization, Writing-Original draft preparation. WSH: Investigation, Conceptualization, Data curation, Methodology, Supervision, Writing-Reviewing and Editing. NT: Validation, Data curation, Writing - Review & Editing. JW: Validation, Data curation. All authors have read and agreed to the submitted version of the manuscript.

## Supplemental Material

**Supplemental Table 1:**
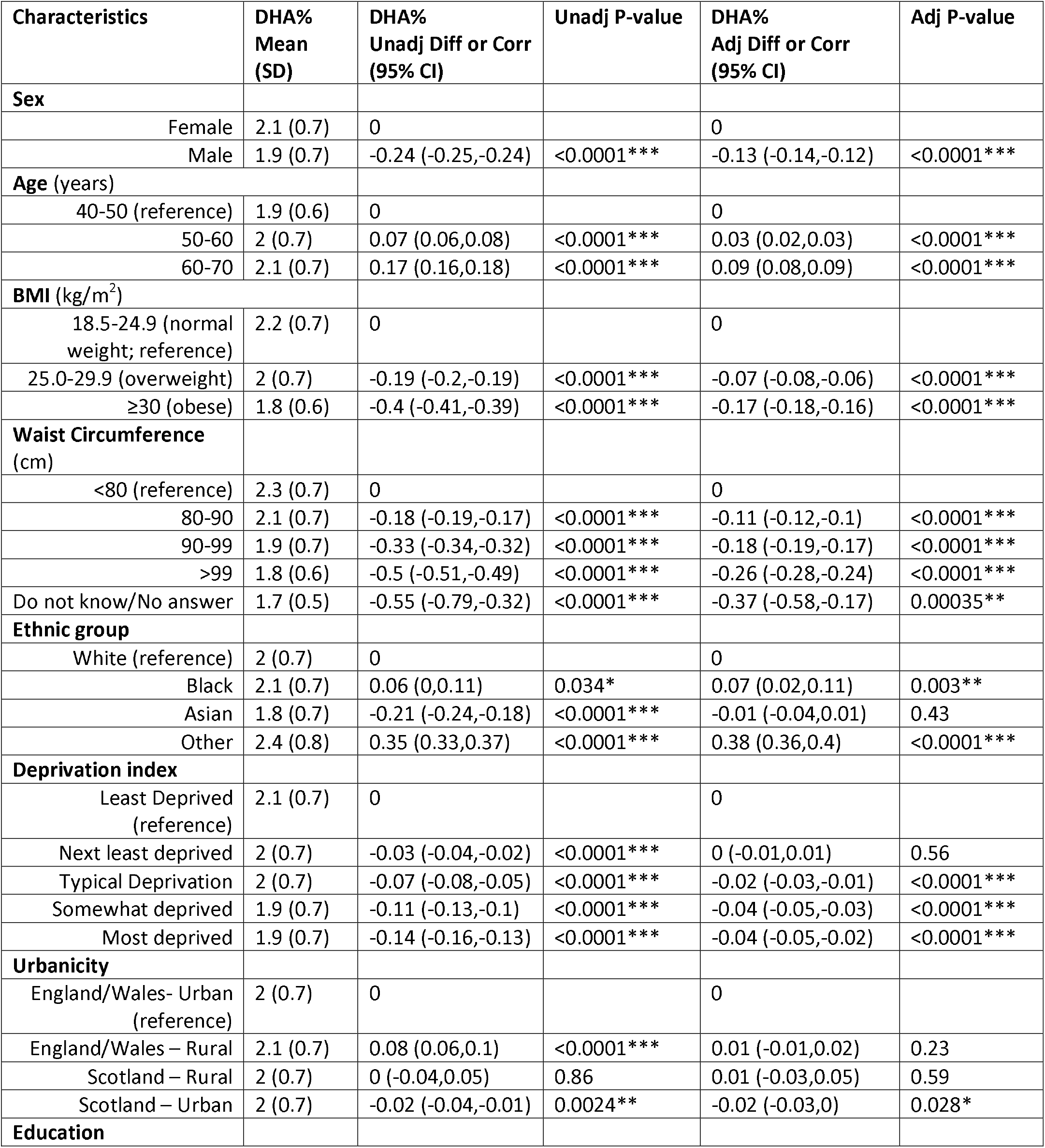
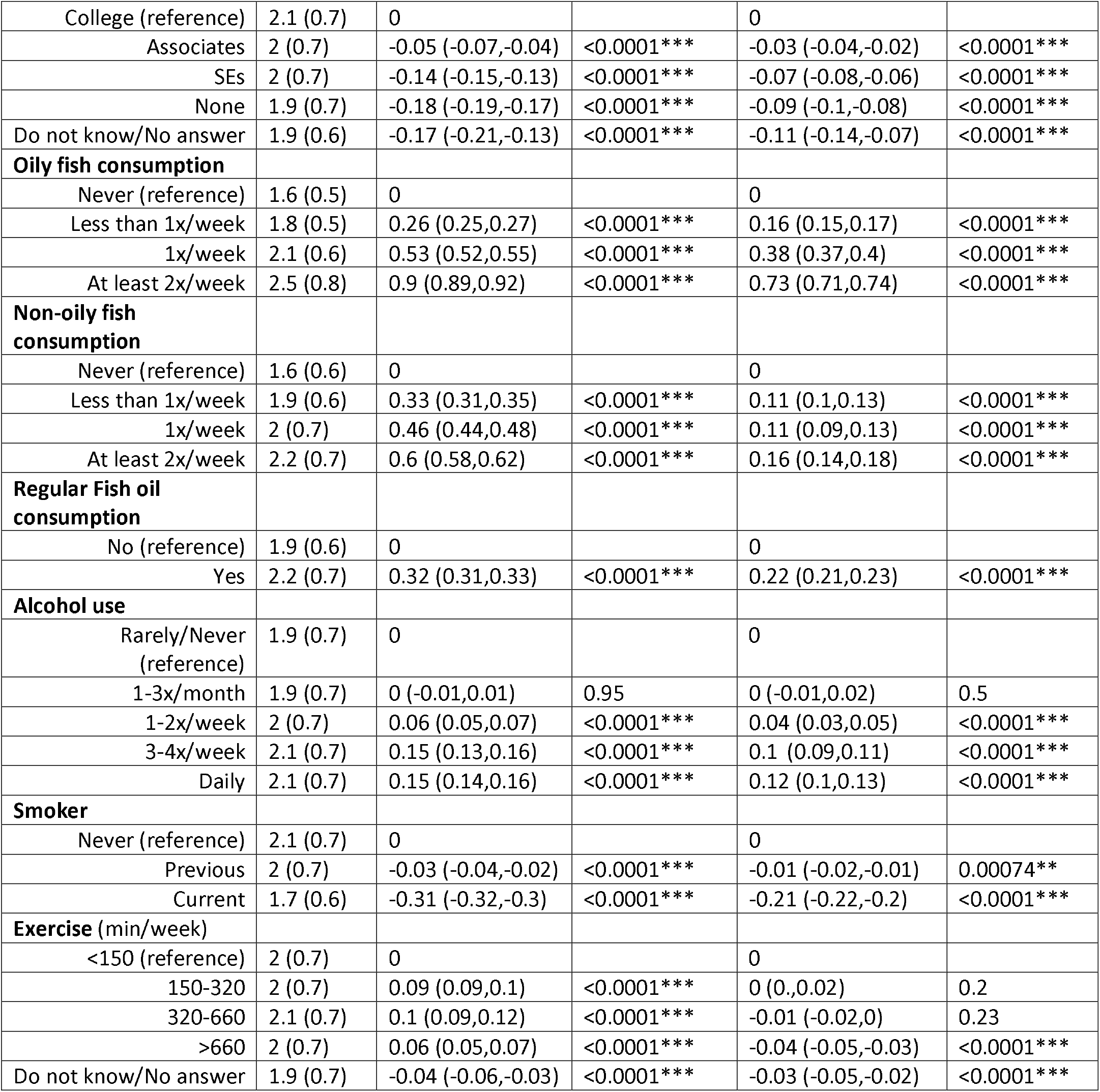
Associations of demographic characteristics and DHA% (measured by NMR) in unadjusted and adjusted (for all variables in Table 1) analyses (n=117,108).

**Supplemental Table 2:** R-squared values for the participant’s characteristics on the variability of DHA% measure by NMR.

|  |  |  |
| --- | --- | --- |
| **Overall model** | 29.2% |  |
| **Variable dropped<sup>1</sup>** | **New R-squared<sup>1</sup>** | **Uniquely explained variability<sup>2</sup>** |
| **Oily fish consumption** | 20.1% | 9.1% |
| **Regular fish oil consumption** | 27.0% | 2.2% |
| **Ethnic group** | 28.4% | 0.9% |
| **Smoking** | 28.4% | 0.8% |
| **Waist Circumference** | 28.6% | 0.6% |
| **Sex** | 28.6% | 0.6% |
| **Alcohol use** | 28.8% | 0.4% |
| **BMI** | 28.8% | 0.4% |
| **Education** | 29.0% | 0.3% |
| **Age** | 29.0% | 0.2% |
| **Non-oily fish consumption** | 29.0% | 0.2% |
| **Exercise** | 29.1% | 0.1% |
| **Urbanicity** | 29.1% | 0.1% |
| **Deprivation index** | 29.2% | 0.1% |

1. New R-squared provides the model R-squared for the other 13 variables when predicting DHA%. Variables are ranked according to their independent contribution on the total variability.
2. Uniquely explained variability is computed as the difference in the model without the variable compared to the full model with all 14 variables (overall model).

**Supplemental Figure 1:**
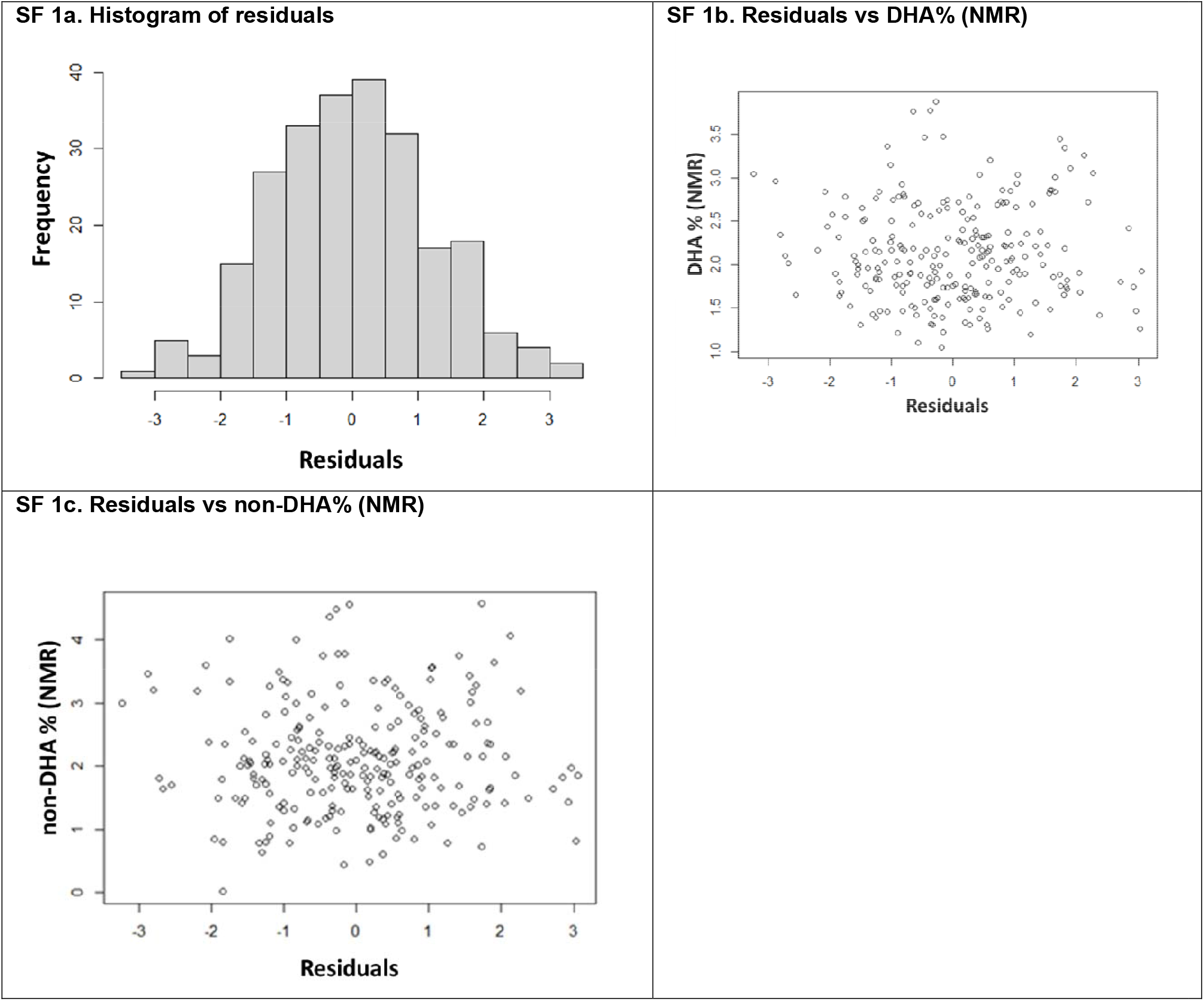
Prediction model diagnostics. Figure 1a shows the normality of the residuals for the final prediction model. Figures 1b and 1c show good model fit characteristics when looking at the prediction model residuals vs both predictor variables (DHA% [Figure 1b]; non-DHA% [Figure 1c]).

## References

1. Hu Y, Hu FB, Manson JE (2019) Marine Omega-3 Supplementation and Cardiovascular Disease: An Updated Meta-Analysis of 13 Randomized Controlled Trials Involving 127 477 Participants. JAHA 8, 19, e013543.

2. Del Gobbo LC, Imamura F, Aslibekyan S et al. (2016) ω-3 Polyunsaturated Fatty Acid Biomarkers and Coronary Heart Disease: Pooling Project of 19 Cohort Studies. JAMA Intern Med 176, 8, 1155–1166.

3. Harris WS, Tintle NL, Imamura F et al. (2021) Blood n-3 fatty acid levels and total and cause-specific mortality from 17 prospective studies. Nat Commun 12, 1, 2329.

4. Liu J, Li X, Hou J et al. (2021) Dietary Intake of N-3 and N-6 Polyunsaturated Fatty Acids and Risk of Cancer: Meta-Analysis of Data from 32 Studies. Nutr Cancer 73, 6, 901–913.

5. Ma M-Y, Li K-L, Zheng H et al. (2021) Omega-3 index and type 2 diabetes: Systematic review and meta-analysis. Prostaglandins Leukot Essent Fatty Acids 174, 102361.

6. Qian F, Ardisson Korat AV, Imamura F et al. (2021) n-3 Fatty Acid Biomarkers and Incident Type 2 Diabetes: An Individual Participant-Level Pooling Project of 20 Prospective Cohort Studies. Diabetes Care 44, 5, 1133–1142.

7. Liu R, Chen L, Wang Y et al. (2020) High ratio of ω-3/ω-6 polyunsaturated fatty acids targets mTORC1 to prevent high-fat diet-induced metabolic syndrome and mitochondrial dysfunction in mice. J Nutr Biochem 79, 108330.

8. Kosti RI, Kasdagli MI, Kyrozis A et al. (2022) Fish intake, n-3 fatty acid body status, and risk of cognitive decline: a systematic review and a dose-response meta-analysis of observational and experimental studies. Nutr Rev 80, 6, 1445–1458.

9. Deyama S, Ishikawa Y, Yoshikawa K et al. (2017) Resolvin D1 and D2 Reverse Lipopolysaccharide-Induced Depression-Like Behaviors Through the mTORC1 Signaling Pathway. Int J Neuropsychopharmacol 20, 7, 575–584.

10. Harris WS & Thomas RM (2010) Biological variability of blood omega-3 biomarkers. Clin Biochem 43, 3, 338–340.

11. Fenton JI, Gurzell EA, Davidson EA et al. (2016) Red blood cell PUFAs reflect the phospholipid PUFA composition of major organs. Prostaglandins Leukot Essent Fatty Acids 112, 12–23.

12. Harris WS & Schacky C von (2004) The Omega-3 Index: a new risk factor for death from coronary heart disease? Prev Med 39, 1, 212–220.

13. Schuchardt JP, Cerrato M, Ceseri M et al. (2022) Red blood cell fatty acid patterns from 7 countries: Focus on the Omega-3 index. Prostaglandins Leukot Essent Fatty Acids 179, 102418.

14. Demonty I, Langlois K, Greene-Finestone LS et al. (2021) Proportions of long-chain ω-3 fatty acids in erythrocyte membranes of Canadian adults: Results from the Canadian Health Measures Survey 2012-2015. Am J Clin Nutr 113, 4, 993–1008.

15. Collins R (2012) What makes UK Biobank special? The Lancet 379, 9822, 1173–1174.

16. Sudlow C, Gallacher J, Allen N et al. (2015) UK biobank: an open access resource for identifying the causes of a wide range of complex diseases of middle and old age. PLoS Med 12, 3, e1001779.

17. Würtz P, Raiko JR, Magnussen CG et al. (2012) High-throughput quantification of circulating metabolites improves prediction of subclinical atherosclerosis. Eur Heart J 33, 18, 2307–2316.

18. DeFina LF, Bassett MH, Finley CE et al. (2016) Association between omega-3 fatty acids and serum prostate-specific antigen. Nutr Cancer 68, 1, 58–62.

19. Würtz P, Havulinna AS, Soininen P et al. (2015) Metabolite Profiling and Cardiovascular Event Risk. Circulation 131, 9, 774–785.

20. Rubin DB (1987) Multiple Imputation for Nonresponse in Surveys. Hoboken, NJ, USA: John Wiley & Sons, Inc.

21. Mackenbach JP (1988) Health and deprivation. Inequality and the North. Health Policy 10, 2, 207.

22. Office for National Statistics, National Records of Scotland, Northern Ireland Statistics and Research Agency (2017) 2011 Census aggegate data (Data downloaded: 1 February 2017).

23. Stark KD, van Elswyk ME, Higgins MR et al. (2016) Global survey of the omega-3 fatty acids, docosahexaenoic acid and eicosapentaenoic acid in the blood stream of healthy adults. Prog Lipid Res 63, 132–152.

24. Block RC, Harris WS, Pottala JV (2008) Determinants of Blood Cell Omega-3 Fatty Acid Content. Open Biomark J 1, 1–6.

25. Flock MR, Skulas-Ray AC, Harris WS et al. (2013) Determinants of Erythrocyte Omega-3 Fatty Acid Content in Response to Fish Oil Supplementation: A Dose–Response Randomized Controlled Trial. JAHA 2, 6.

26. Lorgeril M de, Salen P, Martin J-L et al. (2008) Interactions of wine drinking with omega-3 fatty acids in patients with coronary heart disease: a fish-like effect of moderate wine drinking. Am Heart J 155, 1, 175–181.

27. Groot RHM de, van Boxtel MPJ, Schiepers OJG et al. (2009) Age dependence of plasma phospholipid fatty acid levels: potential role of linoleic acid in the age-associated increase in docosahexaenoic acid and eicosapentaenoic acid concentrations. Br J Nutr 102, 7, 1058– 1064.

28. Harris WS, Pottala JV, Lacey SM et al. (2012) Clinical correlates and heritability of erythrocyte eicosapentaenoic and docosahexaenoic acid content in the Framingham Heart Study. Atherosclerosis 225, 2, 425–431.

29. Itomura M., Fujioka S., Hamazaki K. et al. (2008) Factors influencing EPA + DHA levels in red blood cells in Japan. In Vivo 22, 131–135.

30. Harris WS, Tintle NL, Manson JE et al. (2021) Effects of menopausal hormone therapy on erythrocyte n-3 and n-6 PUFA concentrations in the Women’s Health Initiative randomized trial. Am J Clin Nutr 113, 6, 1700–1706.

31. Burdge GC & Wootton SA (2003) Conversion of α-linolenic acid to palmitic, palmitoleic, stearic and oleic acids in men and women. Prostaglandins, Leukotrienes and Essential Fatty Acids 69, 4, 283–290.

32. Giltay EJ, Gooren LJG, Toorians AWFT et al. (2004) Docosahexaenoic acid concentrations are higher in women than in men because of estrogenic effects. Am J Clin Nutr 80, 5, 1167–1174.

33. Burdge GC, Jones AE, Wootton SA (2002) Eicosapentaenoic and docosapentaenoic acids are the principal products of alpha-linolenic acid metabolism in young men*. Br J Nutr 88, 4, 355–363.

34. Hodson L, Skeaff CM, Fielding BA (2008) Fatty acid composition of adipose tissue and blood in humans and its use as a biomarker of dietary intake. Prog Lipid Res 47, 5, 348– 380.

35. Kalsbeek A, Veenstra J, Westra J et al. (2018) A genome-wide association study of red-blood cell fatty acids and ratios incorporating dietary covariates: Framingham Heart Study Offspring Cohort. PLoS One 13, 4, e0194882.

36. Wagner A, Simon C, Morio B et al. (2015) Omega-3 index levels and associated factors in a middle-aged French population: the MONA LISA-NUT Study. Eur J Clin Nutr 69, 4, 436– 441.

37. Plourde M, Chouinard-Watkins R, Vandal M et al. (2011) Plasma incorporation, apparent retroconversion and β-oxidation of 13C-docosahexaenoic acid in the elderly. Nutr Metab (Lond) 8, 5.

38. Sands SA, Reid KJ, Windsor SL et al. (2005) The impact of age, body mass index, and fish intake on the EPA and DHA content of human erythrocytes. Lipids 40, 4, 343–347.

39. Nevill AM, Duncan MJ, Myers T (2022) BMI is dead; long live waist-circumference indices: But which index should we choose to predict cardio-metabolic risk? Nutr Metab Cardiovasc Dis 32, 7, 1642–1650.

40. Cazzola R, Rondanelli M, Russo-Volpe S et al. (2004) Decreased membrane fluidity and altered susceptibility to peroxidation and lipid composition in overweight and obese female erythrocytes. J Lipid Res 45, 10, 1846–1851.

41. Wang Y, Botolin D, Xu J et al. (2006) Regulation of hepatic fatty acid elongase and desaturase expression in diabetes and obesity. J Lipid Res 47, 9, 2028–2041.

42. Zehr KR, Segovia A, Shah M et al. (2019) Associations of medium and long chain omega-3 polyunsaturated fatty acids with blood pressure in Hispanic and non-Hispanic smokers and nonsmokers. Prostaglandins Leukot Essent Fatty Acids 144, 10–15.

43. Hoge A, Bernardy F, Donneau A-F et al. (2018) Low omega-3 index values and monounsaturated fatty acid levels in early pregnancy: an analysis of maternal erythrocytes fatty acids. Lipids Health Dis 17, 1, 63.

44. Sala-Vila A, Harris WS, Cofán M et al. (2011) Determinants of the omega-3 index in a Mediterranean population at increased risk for CHD. Br J Nutr 106, 3, 425–431.

45. Polidori MC, Mecocci P, Stahl W et al. (2003) Cigarette smoking cessation increases plasma levels of several antioxidant micronutrients and improves resistance towards oxidative challenge. Br J Nutr 90, 1, 147–150.

46. Morrow JD, Frei B, Longmire AW et al. (1995) Increase in circulating products of lipid peroxidation (F2-isoprostanes) in smokers. Smoking as a cause of oxidative damage. N Engl J Med 332, 18, 1198–1203.

47. Di Giuseppe R, Lorgeril M de, Salen P et al. (2009) Alcohol consumption and n-3 polyunsaturated fatty acids in healthy men and women from 3 European populations. Am J Clin Nutr 89, 1, 354–362.

48. Schacky C von, Kemper M, Haslbauer R et al. (2014) Low Omega-3 Index in 106 German elite winter endurance athletes: a pilot study. Int J Sport Nutr Exerc Metab 24, 5, 559–564.

49. Ritz PP, Rogers MB, Zabinsky JS et al. (2020) Dietary and Biological Assessment of the Omega-3 Status of Collegiate Athletes: A Cross-Sectional Analysis. PLoS One 15, 4, e0228834.

50. Moyers B, Farzaneh-Far R, Harris WS et al. (2011) Relation of whole blood n-3 fatty acid levels to exercise parameters in patients with stable coronary artery disease (from the heart and soul study). Am J Cardiol 107, 8, 1149–1154.

51. Cutler DM & Lleras-Muney A (2010) Understanding differences in health behaviors by education. J Health Econ 29, 1, 1–28.

52. Heuer T, Krems C, Moon K et al. (2015) Food consumption of adults in Germany: results of the German National Nutrition Survey II based on diet history interviews. Br J Nutr 113, 10, 1603–1614.

53. Dijkstra SC, Neter JE, Brouwer IA et al. (2014) Adherence to dietary guidelines for fruit, vegetables and fish among older Dutch adults; the role of education, income and job prestige. J Nutr Health Aging 18, 2, 115–121.

54. Li Z-H, Zhong W-F, Liu S et al. (2020) Associations of habitual fish oil supplementation with cardiovascular outcomes and all cause mortality: evidence from a large population based cohort study. BMJ 368, m456.

